# Epidemiological characteristics of COVID-19 patients in Samarinda, East Kalimantan, Indonesia

**DOI:** 10.1101/2020.07.10.20151175

**Authors:** Swandari Paramita, Ronny Isnuwardana, Anton Rahmadi, Osa Rafshodia, Ismid Kusasih

## Abstract

**Introduction:** Coronavirus Disease (COVID-19) is caused by SARS-CoV-2 infection. Indonesia announced the first COVID-19 case on 2 March 2020. East Kalimantan has been determined as the new capital of Indonesia since 2019. This makes Samarinda as the capital of East Kalimantan has been focused for its capability of handling COVID-19 patients. We report the epidemiological characteristics and immunofluorescence assay results of these patients.

**Methods:** All patients with positive confirmed COVID-19 by RT-PCR were admitted to hospitals and quarantine center in Samarinda. We retrospectively analyzed data from the daily report of the Samarinda City and East Kalimantan Health Office information system.

**Results:** By June 25, 2020, 64 patients had been identified as having positive confirmed COVID-19. The mean age of the patients was 37.3 ± 13.8 years. Most of the patients were men (57 [90.6%] patients). Thirty-nine COVID-19 patients were imported cases with a history of traveling from South Sulawesi. Most of the patients were admitted to the Quarantine Center of Samarinda City. The mean duration from the first hospital admission for isolation to discharge was 25.6 ± 13.1 days. There was only one death case of COVID-19 patients in Samarinda. There were the highest confirmed cases of COVID-19 in Samarinda in early June 2020. There was a declining trend in the age of COVID-19 patients and the duration of isolation time in the hospital.

**Discussion:** Imported cases still contributed to the increase of COVID-19 cases in Samarinda. Younger age of COVID-19 patients was more involved in frequent mobility which makes them cause the spread of the disease. Activation of the national reference laboratory for the COVID-19 examination in Samarinda has reduced the length of time patients treated in hospitals.

**Conclusion:** The epidemiological characteristics of COVID-19 patients show the ability of local governments to deal with this pandemic. This can be seen from the low case fatality rate in Samarinda.

## Introduction

Coronavirus Disease (COVID-19) is caused by SARS-CoV-2 infection. Coronavirus is an RNA virus from the Coronaviridae family (1). Although most coronavirus infections are mild, there have been two epidemics of coronavirus in the past, namely severe acute respiratory coronavirus syndrome (SARS) (2) and Middle East respiratory syndrome coronavirus (MERS) (3), with a mortality rate of 10% for SARS (4) and 37% for MERS (5).

A series of pneumonia cases emerged in Wuhan, China, in December 2019, with clinical findings of viral pneumonia. (6). The analysis of patient samples indicates a new coronavirus, which was then named SARS-CoV-2 (7). Until the end of June 2020, the USA, Brazil, and Russia were the countries for the most COVID-19 cases in the world. There are more than 10 million positive confirmed cases worldwide (8).

On March 2, 2020, Indonesia announced the first case of COVID-19 (9). On March 18, 2020, the first case of COVID-19 in East Kalimantan was a patient from Samarinda (10). East Kalimantan province has been determined as the new capital of Indonesia by 2019 (11). This makes Samarinda as the capital of East Kalimantan has been focused for its capability of handling COVID-19 patients (12). Therefore, the study aims to describe the epidemiological characteristics of COVID-19 patients in Samarinda.

## Methods

### Study Design

This study was a descriptive analysis of all cases of COVID-19 diagnosed in Samarinda, East Kalimantan, Indonesia as of the end of June 25, 2020. This study was approved by the Ethical Health Research Commission of Faculty of Medicine Mulawarman University, Samarinda, Indonesia. Although individual informed consent was not required for this study, all data were handled to protect patient confidentiality and privacy.

### Data Source

All data contained in all COVID-19 case records in the Samarinda City and East Kalimantan Health Office information system through the end of June 25, 2020, were extracted from the system. No sampling was done to achieve a predetermined study size and no eligibility criteria were used (all cases were included).

### Variables

Patient characteristics were collected at the time of diagnosis and entry into the Samarinda City and East Kalimantan Health Office information system. All confirmed cases were diagnosed based on examination to confirm COVID-19 using reverse transcription-polymerase chain reaction (RT-PCR) according to guidelines from the Ministry of Health of the Republic of Indonesia. Some of the patients going through immunofluorescence assay (IFA) testing for immunoglobulin (Ig) G and M level.

### Analysis

For all cases, epidemiological characteristics were summarized using descriptive statistics. Continuous variables were expressed as a mean and standard deviation; categorical variables were expressed as number (%). Statistical analyses were done using Microsoft Excel.

## Results

By June 25, 2020, 64 patients had been identified as having laboratory-confirmed COVID-19. Twenty-three (35.4%) of the COVID-19 patients were aged 30-39 years. The mean age of the patients was 38.4 ± 13.4 years. Most of the infected patients were men (59 [90.8%]). Thirty-nine (60%) of the COVID-19 patients in Samarinda were imported cases with a history of traveling from South Sulawesi. Most of the patients were admitted to Quarantine Center Samarinda City (39 [60%]). The mean duration from COVID-19 laboratory confirmation to patient recovery was 15.2 ± 10.5 days. Patients with clinical improvement can be discharged from the hospital if the results of RT-PCR examination two days in a row show negative results. The case fatality rate was 0.02. There was only one death case of COVID-19 patient in Samarinda until June 25, 2020.

**Table 1.** Epidemiological characteristics of COVID-19 patients.

| Characteristics | Mean ± SD or N (%) |
| --- | --- |
| Age, years | 37.3 ± 13.8 |
| <10 years | 2 (3.1%) |
| 10-19 years | 1 (1.6%) |
| 20-29 years | 16 (25.0%) |
| 30-39 years | 21 (32.8%) |
| 40-49 years | 12 (18.8%) |
| 50-59 years | 6 (9.4%) |
| 60-69 years | 5 (7.8%) |
| >69 years | 1 (1.6%) |
| Sex |  |
| Men | 57 (90.6%) |
| Women | 7 (10.9%) |
| Mode of transmission imported cases |  |
| From South Sulawesi | 39 (60.9%) |
| From East Java | 7 (10.9%) |
| From Jakarta | 4 (6.3%) |
| From Central Kalimantan | 2 (3.1%) |
| From South Kalimantan | 2 (3.1%) |
| From North Maluku | 1 (1.6%) |
| From Regencies in East Kalimantan | 9 (14.1%) |
| Hospital admission of patients |  |
| AW Sjahrane Hospital (Province Hospital) | 16 (25.0%) |
| IA Moeis Hospital (City Hospital) | 4 (6.3%) |
| Quarantine Center of Samarinda City | 39 (60.9%) |
| Others | 5 (7.8%) |
| Case Fatality Rate (CFR) | 0.02 |
| Days from isolation to discharge | 25.6 ± 13.1 |
| RT-PCR testing | 64 (100%) |
| IFA testing | 47 (73.4%) |
| IgG (IU/mL) | 20.0 ± 13.6 |
| IgM (IU/mL) | 0.7 ± 1.0 |

On the 12th week (June 3-9, 2020) there were the highest confirmed cases reaching 1.9 patients per day. This number has gradually declined in the following weeks (Figure 1). There is a declining trend in the age of COVID-19 patients, who are initially over 40 years old, later in their 30s (Figure 2). There is also a declining trend in the duration of admission time in the hospital, which initially can last more than 3 weeks, later to be around one week only (Figure 3).

**Figure 1.**
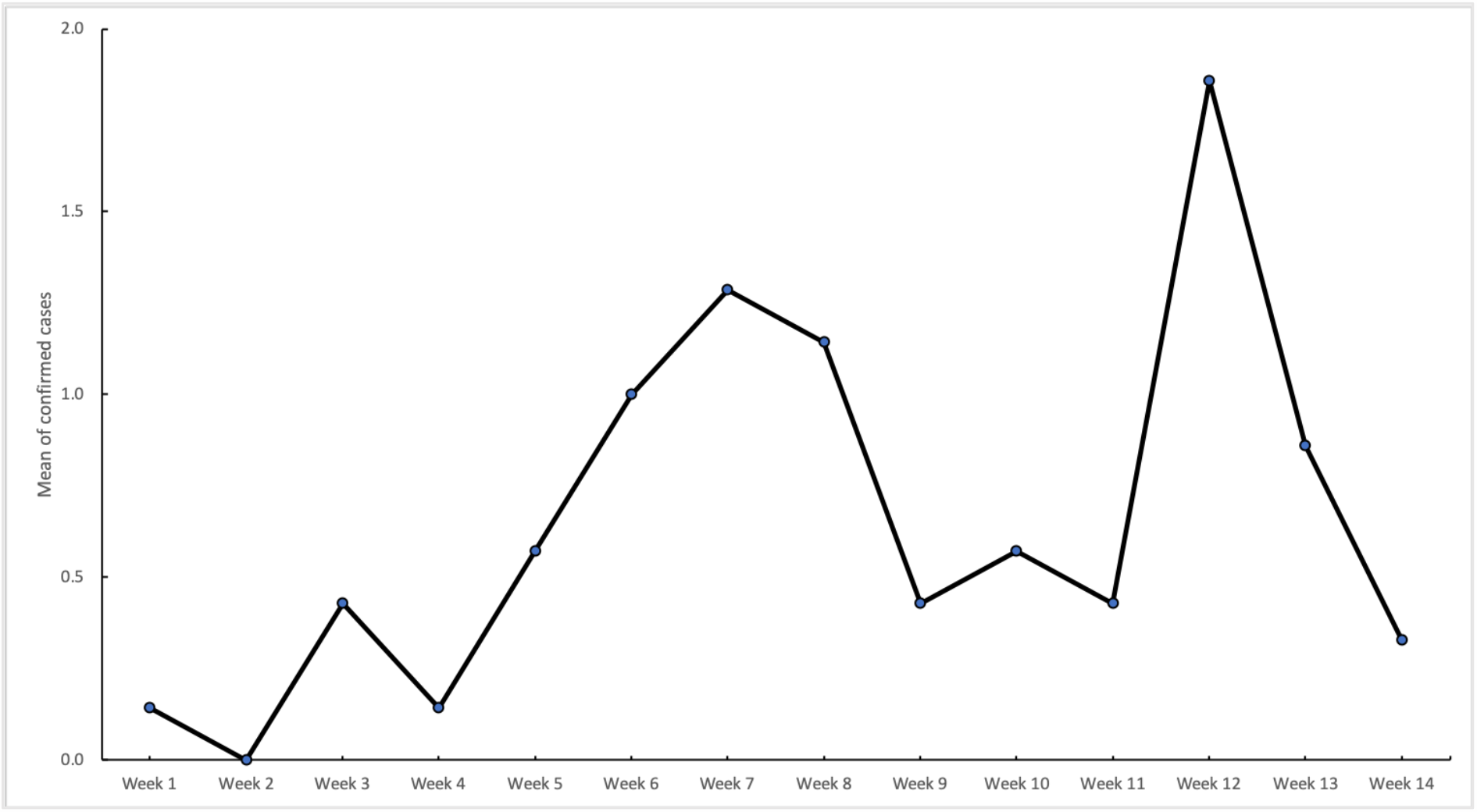
The weekly mean for confirmed cases of COVID-19 in Samarinda (March 18 to June 25, 2020)

**Figure 2.**
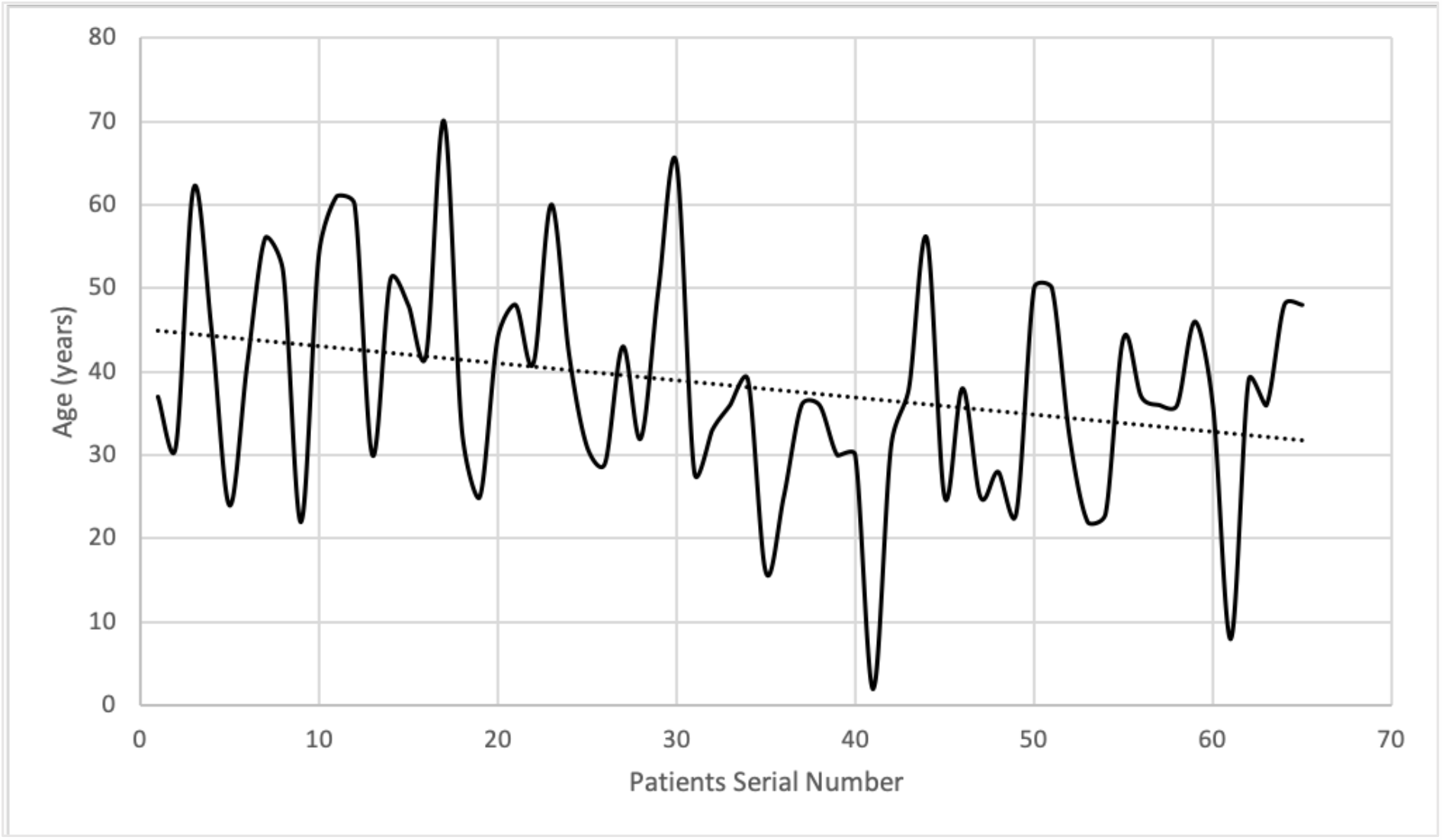
Age trend for confirmed cases of COVID-19 in Samarinda.

**Figure 3.**
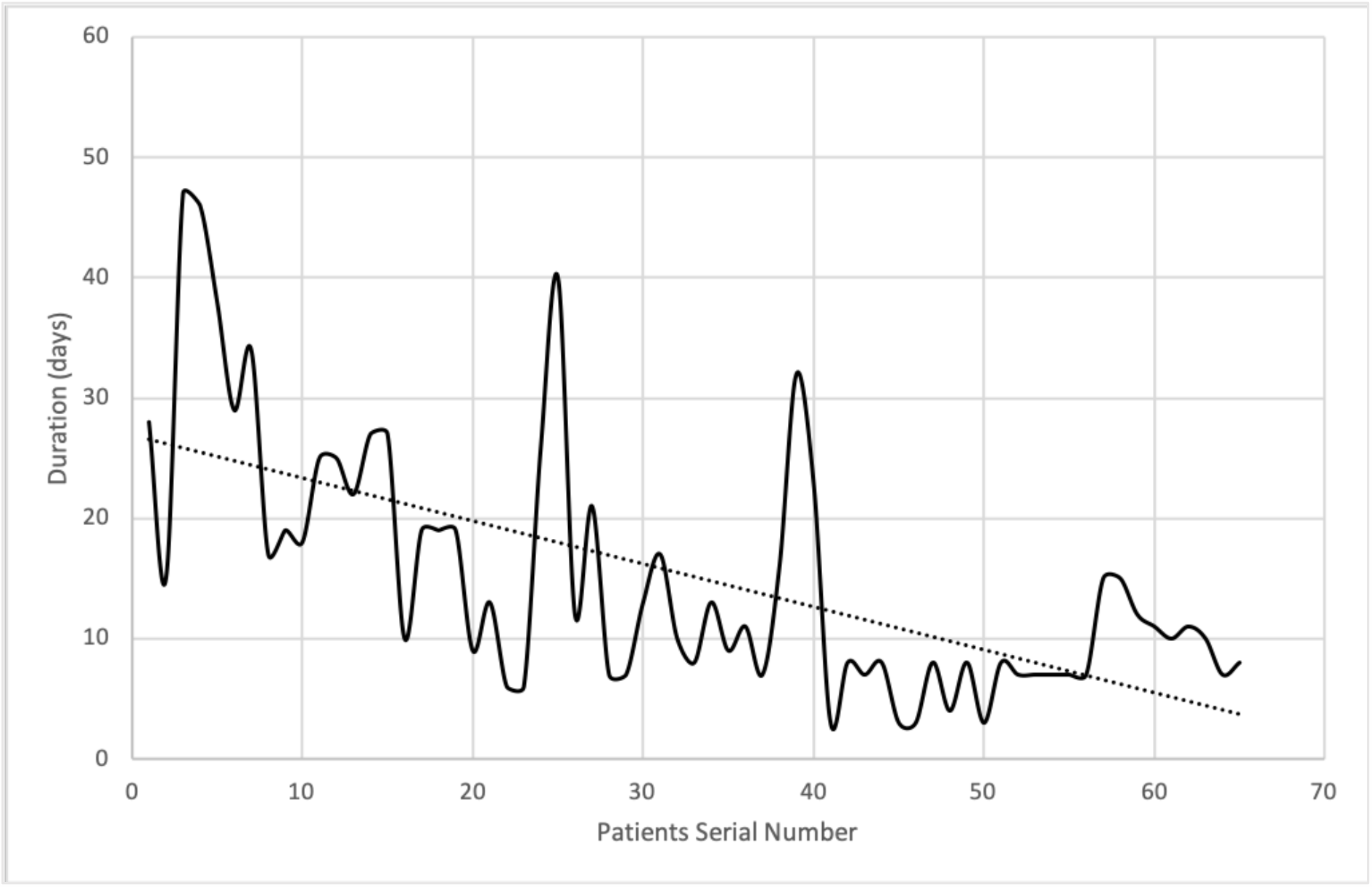
Duration of hospital admission trend for confirmed cases of COVID-19 in Samarinda.

## Discussion

This study reports a study of 64 patients with positive COVID-19 in Samarinda, East Kalimantan, Indonesia. As of June 25, 2020, 100 days after the first confirmed case in Samarinda, there have been 473 positive COVID-19 positive cases in East Kalimantan (13). To date, there are 50,178 positive confirmed cases of COVID-19, with 20,449 people recovering and 2,620 patients have died in Indonesia (14). Most of the infected patients were aged 30-39 years. These results are similar to previous studies at Abdul Wahab Sjahranie Hospital Samarinda which showed that most patients were also in this age group (15). There is a declining trend for the younger age of COVID-19 patients. Young adults are more involved in frequent social activity and mobility which makes them cause the spread of COVID-19 (16). The majority of COVID-19 patients in Samarinda are male. This is consistent with a meta-analysis study which showed that men took the largest percentage in the distribution of COVID-19 according to gender (17).

Sixty percent of the COVID-19 patients in Samarinda were imported cases with a history of traveling from South Sulawesi. Travel history of attending religious activities in Gowa, South Sulawesi a few months ago contributed greatly to the positive confirmed case in Samarinda (18). The cluster of ship crews and shipping agents from South Sulawesi also added the number of imported cases from there. South Sulawesi is one of the three new epicenter COVID-19 in Indonesia, along with South Kalimantan and East Java. The emergence of this new epicenter occurred in early June 2020, together with the time of the highest confirmed COVID-19 cases in Samarinda on the 12th week between 3-9 June 2020 (19).

Most of the patients were admitted to the Quarantine Center of Samarinda City. As of June 25, 2020, only 1 fatal case of confirmed COVID-19 was reported in Samarinda, with CFR 0.02 (13). Most of the COVID-19 patients admitted to hospitals in Samarinda are in mild condition. Quarantine center was established to treat people without symptoms, so the referral hospital will not be full of COVID-19 patients and focus on treating patients with more serious conditions (20).

The average length of laboratory positive COVID-19 to hospital discharge is more than 15 days. This is because COVID-19 patients with clinical improvement can only be discharged from the hospital if the results of RT-PCR examination two days in a row show negative results (21). Before the end of May 2020, the nasopharyngeal swab sample from East Kalimantan being sent to the Center for Health Laboratory of the Ministry of Health in Surabaya, Indonesia. This laboratory serves the examination of reference specimens from the provinces of East, Central, South, and North Kalimantan (12). The large number of samples that have to be examined up to 4 provinces makes the slow progress of laboratory examination. Patients must wait a long time for laboratory confirmation results of COVID-19 (22). From May 21, 2020, there are 3 national reference laboratories for the COVID-19 examination in East Kalimantan, namely Abdul Wahab Sjahranie Hospital Samarinda, Kanujoso Djatiwibowo, and Pertamina Hospital Balikpapan (23). Since then the length of time patients have been hospitalized has been reduced from the initial 3 weeks to one week.

## Conclusion

Imported cases still contributed to the increase of COVID-19 cases in Samarinda. Younger age of COVID-19 patients was more involved in frequent mobility which makes them cause the spread of COVID-19. Activation of the national reference laboratory for the COVID-19 examination in Samarinda has reduced the length of time patients treated in hospitals. The epidemiological characteristics of COVID-19 patients show the ability of local governments to deal with this pandemic. This can be seen from the low case fatality rate in Samarinda.

## Data Availability

Data were available on request from the authors.

